# Impaired lung function is associated with elevated blood biomarkers of AD/ADRD: Unraveling the interplay with risk of dementia

**DOI:** 10.1101/2025.05.29.25328529

**Authors:** Sithara Vivek, Eileen M Crimmins, Jung Ki Kim, Jessica Faul, David R Jacobs, Weihua Guan, Bharat Thyagarajan

**Affiliations:** Department of Laboratory Medicine and Pathology, University of Minnesota, Minneapolis, MN, United States; Leonard Davis School of Gerontology, University of Southern California, Los Angeles, CA, United States; Survey Research Center, Institute for Social Research, University of Michigan, Ann Arbor, MI, United States; Division of Epidemiology and Community Health, School of Public Health, University of Minnesota, United States; Division of Biostatistics, School of Public Health, University of Minnesota, Minneapolis MN, United States

## Abstract

**Rationale:** We hypothesize that impaired lung function contributes to dementia risk via biological mechanisms reflected in AD/ADRD-related protein biomarkers, and aim to evaluate their role in mediating this relationship.

**Methods:** Serum p-Tau181 and plasma Aβ42/40 ratio, NfL, and GFAP were measured in 4,072 participants (mean age 66 ± 10; 59% women) in the 2016 Health and Retirement Study. Peak Expiratory Flow (PEF) was assessed in 2012/2014, and cognitive function was measured biennially from 2014–2020 to determine dementia status. Impaired lung function (ILF) was defined as PEF <80% predicted. Multivariable regression examined associations between lung function and AD biomarkers; causal mediation analysis evaluated their role in linking lung function to incident dementia.

**Results:** In total, 881 (21.6%) participants had ILF and 272 (6.8%) participants developed dementia. After adjusting for demographics, education, BMI, smoking, comorbidities, inflammation, eGFR and *APOE e4*, ILF was associated with a higher risk of dementia (HR=1.74; 95% CI (1.34, 225)). Individuals with ILF had 0.10 SD higher NfL (SE= 0.03; p= 0.004) and 0.09 SD higher p-Tau 181 (SE= 0.03; p= 0.002) compared to those without ILF. NfL mediated 7.3% (p=0.01) of the total effect of ILF on dementia, while p-Tau 181 mediated 5% (p=0.05) of this association.

**Conclusions:** ILF was associated with elevated levels of neurodegeneration markers NfL and p-Tau 181, which partially mediated its relationship with dementia risk. These findings highlight the importance of monitoring blood protein biomarkers in individuals with impaired lung health to facilitate early interventions.

## INTRODUCTION

Extensive research across diverse populations has consistently shown a link between impaired lung health (ILF) and poor cognitive outcomes^1–5^, suggesting that maintaining optimal lung function may play a critical role in preserving cognitive abilities. Multiple cohort studies have shown that reduced lung function is associated with cognitive decline and an increased risk of dementia^5–7^. Studies with extended follow-up data reported that improved lung function measures - such as FEV1, FVC and FEV1/FVC ratio - were associated with a slower rate of cognitive decline across multiple domains, including memory, language, and processing speed/attention^4,8^. A study in in the Atherosclerosis Risk in Communities (ARIC) cohort showed that lung disease, including both restrictive and obstructive lung diseases, among middle-aged adults was associated with increased risk of developing dementia or cognitive impairment later in life^9^. Additionally, a recent study in the National Health and Aging Trends Study (NHATS) cohort, a nationally representative sample of older adults in the US, found that higher levels of lung function measured by peak expiratory flow (PEF) was associated with lower risk of developing dementia, exhibiting a dose-dependent relationship^3^. There has been some effort in understanding the mechanisms linking ILF and poor cognitive outcomes. A study in the Rush Memory and Aging Project demonstrated a link between poor pulmonary function and pathological features of Alzheimer’s disease (including global AD pathology, amyloid beta [Aβ] load, and neurofibrillary tangles) and cerebral vascular disease pathology (including atherosclerosis and cerebral amyloid angiopathy). These findings suggest that both neurodegenerative and vascular mechanisms may play a role in linking ILF and dementia^10^. Moreover, recent meta-analyses^11^ have shown associations between ILF and brain imaging biomarkers of neurodegeneration, vascular and AD pathology. Despite these findings, there remains a significant gap in our understanding of the molecular mechanisms linking impaired lung function to a higher risk of dementia. Identifying these molecular mechanisms is essential for understanding how impaired lung function contributes to neurodegeneration and cognitive decline.

Recent advancements in highly sensitive protein assays enable measurement of low protein levels using minimal blood samples. These less invasive blood-based protein biomarkers have shown promise in early identification of cognitive decline and in staging of Alzheimer’s disease (AD) and AD-related dementias (ADRD), offering improved utility in population studies that support research into AD pathogenesis^12–14^. Several key protein biomarkers measured in plasma and serum, including Amyloid beta 42/40 ratio^15^, Neurofilament Light Chain (NfL)^16^, Glial Fibrillary Acidic Protein (GFAP), and phosphorylated tau (p-Tau 181 and 217^12,17^) were associated with decline in cognitive function and risk of ADRD^16^. Plasma p-Tau proteins have emerged as a promising candidate marker during symptomatic and preclinical AD when it is used with Aβ42/Aβ40^13^. A recent case-control study demonstrated the promise of using all plasma biomarkers and *APOE e4* for prediction of AD clinical diagnosis that reached area under receiver operating characteristic curve (AUC) = 0.81^18^. Increasing research in blood-based biomarkers has demonstrated the clinical utility of these AD protein biomarkers for risk stratification and targeted interventions^13,19^. Increasing the validity of biomarker measures in AD/ADRD is crucial for improving early diagnosis and potentially developing treatments for this condition. Identifying and accounting for factors that affect biomarker concentrations is an essential step in achieving this goal. A recent study demonstrated the need for accounting for renal function and obesity in the measurement of NfL and GFAP^20^. A study among Multiple Sclerosis patients showed an inverse association between BMI and levels of circulating serum NfL^21^ and GFAP^22^. However, the relationship between lung function and blood biomarkers of AD and their mediating role with cognitive function, are still unclear.

We hypothesized that impaired lung function is associated with higher levels of protein biomarkers of AD/ADRD and these biomarkers of neuropathology mediate the association between impaired lung function to increased risk of dementia in older adults. To test this hypothesis, we aimed (1) to evaluate the association between lung function (PEF) and levels of blood biomarkers of AD (Aβ 42/40 ratio, p-Tau 181, NfL, and GFAP) and, (2) to investigate the role of AD biomarkers mediating the association between impaired lung function (ILF) and risk of dementia in a nationally representative sample of older adults in the Health and Retirement Study (HRS).

## METHODS

### Study design and participants

Health and Retirement Study (HRS) is a biennial survey of older adults in the United States that started in 1992 based on a multi-stage area probability design involving geographical stratification and clustering and oversampling of certain demographic groups and collects a wide-range of data on health, biomarkers, genetics, employment, wealth and family^23^. HRS follows participants longitudinally until death and employs a steady-state design to replenish the sample with new participants to maintain population representativeness as the study sample has aged. Additional details of the HRS study design and measurements can be found in previous publications^24–26^. We analyzed data from a subsample of individuals (n=4427) who participated in the 2016 HRS Venous Blood Study (VBS)^27^ and had AD biomarker assessments. The final analytic sample for the primary analysis comprised of 4072 individuals after excluding those missing data on exposure, outcomes or covariates are shown in Figure 1.

**Figure 1:**
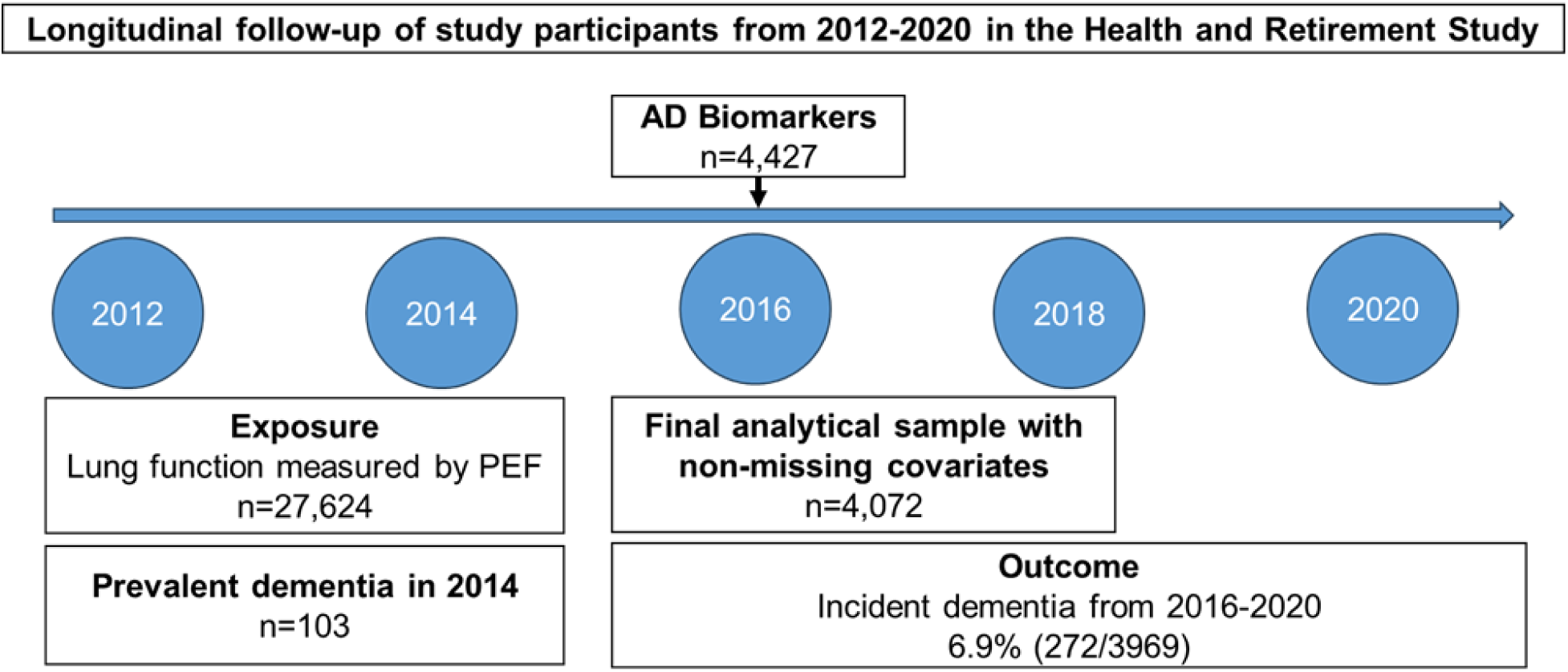
Study design and timeline: In the Health and Retirement Study (HRS) cohort, we identified participants who have lung function measure available in 2012/2014 biennial surveys and blood AD biomarkers measured in 2016 survey and cognitive function measures available from 2014 – 2020 every two years.

The HRS has been approved by the Health Sciences and Behavioral Sciences Institutional Review Board at the University of Michigan. Informed consent was obtained from all respondents in the HRS.

### Exposure measurement: Peak Expiratory Flow (PEF)

In the HRS, trained interviewers employed a standardized assessment of lung function using a peak flow meter, measuring how much air a person can exhale in one breath and reporting the measure as peak expiratory flow (PEF) in L/minute^28^. The assessment was repeated three times. We used the highest of three PEF readings from the 2012 or 2014 HRS in-person visits, as physical measures were collected from a random half-sample in 2012 and the remaining half in 2014 due to the biennial data collection design. Subsequently, we estimated the percent predicted PEF using Hankinson’s equation^29^, which accounts for individual characteristics including age, sex, race and height. We classified participants as having ‘Impaired lung function (ILF)’ if their percent predicted PEF was less than 80% based on the baseline 2012/2014 measure. To assess lung function decline, we calculated the 4-year change in percent predicted PEF from the 2012/2014 to 2016/2018 measures and standardized it using Z-scores.

### Blood-based AD protein biomarkers

AD protein biomarkers were measured in a probability sample drawn from HRS participants in the 2016 Venous Blood Study (VBS). This included individuals aged 60 and older eligible for the 2016 Harmonized Cognitive Assessment Protocol (HCAP), as well as a random half-sample of participants under age 65 who are expected to be eligible for a future HCAP^27^. The Quanterix Simoa Human Neurology 4-Plex E (N4PE) assay was used to measure levels of three biomarkers from plasma samples, amyloid beta 42/ 40 ratio(Aβ42/40), Glial Fibrillary Acidic Protein (GFAP), and Neurofilament light (NfL). Serum was used to assay p-Tau 181. Sample preparation and assays were performed in the Advanced Research Diagnostics Laboratory (ARDL), Minneapolis at the University of Minnesota based on the protocol previously validated^30^.

### Incident dementia

Cognitive function was assessed in the HRS every 2 years from 2014 to 2020. A composite score of overall cognitive performance consisted of scores from four tests: immediate and delayed 10-noun word recall, serial 7-subtraction test, and a backward count from 20. Based on previously published work, we employed the Langa-Weir classification algorithm^24^ to define dementia based on the 27-point cognitive function scale. Participants scoring between 0 and 6 on the 27-point scale were classified as having Dementia, those scoring between 7 and 11 as having Cognitive impairment no dementia (CIND), and those scoring between 12 and 27 as Normal. After excluding participants with dementia in the 2014 survey, we estimated incident dementia among those with Normal or CIND status, using cognitive test scores from the 2016, 2018, and 2020 surveys. Follow-up time ranged from 6 to 8 years, depending on whether lung function was measured in 2014 or 2012, respectively.

### Covariates

Demographic characteristics at baseline—including age, sex, race/ethnicity (White, Black, Hispanic, and Other), smoking status (current, former, or never smoker), and years of education— were collected during the 2012/2014 core survey. Body mass index (BMI) was calculated using measured height and weight from 2012/ 2014 surveys or self-reported values if measured height and weight were not available. BMI (kg/m²) was calculated using the equation weight (pounds)/ (height * height (inches)) * 703. We estimated comorbidity index by counting the number of self-reported chronic conditions such as type 2 diabetes, cancer, hypertension, stroke, heart condition, arthritis and psychiatric problems. We calculated estimated glomerular filtration rate (eGFR) using the new CKD Epi race-free equation based on serum levels of creatinine and cystatin C measures in the 2016 VBS^27^. An inflammatory latent variable was estimated using a confirmatory factor analysis to represent systemic inflammation based on C-reactive protein (*hsCRP*), neutrophil to lymphocyte ratio (NLR) and Cytokines (*IL-6, IL-10, IL-1RA, IGF1,* and *sTNFR-1*) measured in the 2016 VBS^31^. Additionally, we adjusted for *APOE* ε4 allele status, a genetic risk factor for Alzheimer’s disease (AD), determined by the TaqMan assay, with carriers defined as individuals possessing one or two ε4 alleles ^32,33^.

### Statistical analysis

AD biomarkers were log-transformed to address skewed distributions and then standardized to facilitate comparability with other cohorts in statistical analyses. We standardized the % predicted PEF in 2012/2014 and the change in PEF from 2012/2014 to 2016/2018, so that one unit corresponds to one standard deviation. We used ANOVA tests for continuous variables and X2 tests for categorical variables to determine differences in participant characteristics across ILF and normal PEF groups. Multi-variable linear regression models were used to determine the association of baseline ILF (2012/2014) and decline in lung function (from 2012/2014 to 2016/2018) with each continuous measure of AD biomarkers in the 2016 survey. Age, sex, race/ethnicity, years of education, BMI, smoking status and comorbidity index were potential confounding variables in the initial analysis. Additional adjustments for renal function (eGFR), systemic inflammation and *APOE* ε*4* allele status was included in the final statistical models.

We used Cox proportional hazards regression model to estimate association between ILF and risk of dementia and reported hazard ratios and 95% CI over six years of follow-up. For participants who did not develop dementia, follow-up time was censored at their last assessment in the 2020 core survey. Causal mediation analysis was performed to assess the mediating role of AD biomarkers in linking the association between ILF and risk of dementia using the *causalmed* procedure in SAS. All statistical analyses were performed in SAS v9.4 (SAS Institute, Inc., Cary, NC).

## RESULTS

Among 4072 participants in the study, 58.9% were women (n=2397), with mean (±SD) age of 66.2 (±10.3), 21.6% (n=881) had ILF (% predicted PEF < 80%) and 2.5% (n=103) had prevalent dementia in 2014 and 6.9% (n=272) developed dementia over 6 years of follow-up. Black, Hispanic individuals and current smokers had a higher prevalence of ILF at baseline (Table 1).

**Table 1:** Descriptive statistics of participant characteristics in the HRS 2012/2014 survey across lung function groups.

| **Estimates**<br>**Mean±SD / Frequency (%)** | **Overall**<br>**n=4072** | **Normal**<br>**(Predicted PEF**<br>**≥ 80%)**<br>**n=3191 (78.4%)** | **Impaired lung**<br>**function**<br>**(Predicted PEF**<br>**<80%)**<br>**n=881 (21.6%)** | **p value** |
| --- | --- | --- | --- | --- |
| **% predicted PEF 2012/2014** | 98.96 ± 25.74 | 108.65 ± 19.08 | 63.85 ± 13.03 | <.0001 |
| **Age 2014 (years)** | 66.19 ± 10.29 | 66.32 ± 10.26 | 65.70 ± 10.38 | 0.11 |
| **Sex (% females)** | 2397 (58.9%) | 1891 (59.3%) | 506 (57.4%) | 0.3300 |
| **Race/ethnicity** |  |  |  |  |
| Blacks | 667 (16.4%) | 499 (15.6%) | 168 (19.07%) | 0.0003 |
| Hispanics | 601 (14.8%) | 453 (14.2%) | 148 (16.8%) |  |
| Other | 129 (3.2%) | 91 (2.9%) | 38 (4.3%) |  |
| Whites | 2675 (65.7%) | 2148 (67.3%) | 527 (59.8%) |  |
| **Smoking status 2014** |  |  |  |  |
| Current | 501 (12.3%) | 284 (8.9%) | 217 (24.6%) | <.0001 |
| Former | 991 (24.3%) | 790 (24.8%) | 201 (22.8%) |  |
| Never | 2580 (63.4%) | 2117 (66.3%) | 463 (52.6%) |  |
| **Education** |  |  |  |  |
| **0-11 y** | 747 (18.3%) | 503 (15.8%) | 244 (27.7%) | <.0001 |
| **12 y** | 1252 (30.8%) | 944 (29.6%) | 308 (35.0%) |  |
| **13-15y** | 1023 (25.1%) | 843 (26.4%) | 180 (20.4%) |  |
| **16+ y** | 1050(25.8%) | 901 (28.2%) | 149 (16.9%) |  |
| **Body Mass Index (kg/m<sup>2</sup>)**<br>**2012/2014** | 30.56 ± 6.91 | 30.65 ± 6.74 | 30.27 ± 7.50 | 0.1400 |
| **Comorbidity Index 2014** | 1.88 ± 1.29 | 1.83 ± 1.26 | 2.06 ± 1.36 | <.0001 |
| **eGFR (Cys and CR) 2016** | 68.79 ± 21.08 | 68.61 ± 21.17 | 69.44 ± 20.76 | 0.3100 |
| **Inflammatory latent variable**<br>**2016** | 0.08 ± 0.51 | 0.09 ± 0.51 | 0.04 ± 0.49 | 0.0060 |
| **APOE e4 allele, %Yes** | 1122 (27.6%) | 898 (28.1%) | 224 (25.4%) | 0.1100 |
| **Cognitive function 2014 (0-27)** | 15.55 ± 4.22 | 15.89 ± 4.14 | 14.32 ± 4.28 | <.0001 |
| **Cognition category in 2014** |  |  |  | <.0001 |
| Normal | 3388 (83.2%) | 2742 (85.9%) | 646 (73.3%) |  |
| CIND | 581 (14.3%) | 380 (11.9%) | 201 (22.8%) |  |
| Dementia | 103 (2.5%) | 69 (2.2%) | 34 (3.9%) |  |
| **Incident dementia\* n=3969** | 272 (6.9%) | 172 (5.5%) | 99 (11.6%) | <.0001 |

**Note:** \* Dementia based on cognitive scores of 1-6 in 2016, 2018 or 2020 among those who had normal cognition or CIND in 2014.

### Association between lung function and blood biomarkers of AD

We observed that lower baseline percent predicted PEF, modeled as a continuous predictor, was significantly associated with higher levels of p-Tau181, NfL, and GFAP after adjusting for all covariates (Table 2A). No significant association was observed between PEF and the Aβ42/40 ratio. When PEF was modeled in quartiles, individuals in the lowest quartile (Q1) had significantly higher levels of p-Tau181 (β = 0.15, SE = 0.04, p = 0.0002), NfL (β = 0.11, SE = 0.03, p = 0.0006), and GFAP (β = 0.10, SE = 0.03, p = 0.004) compared to those in the highest quartile (Q4), with a clear dose-response gradient across quartiles (Table 2A).

**Figure 2:**
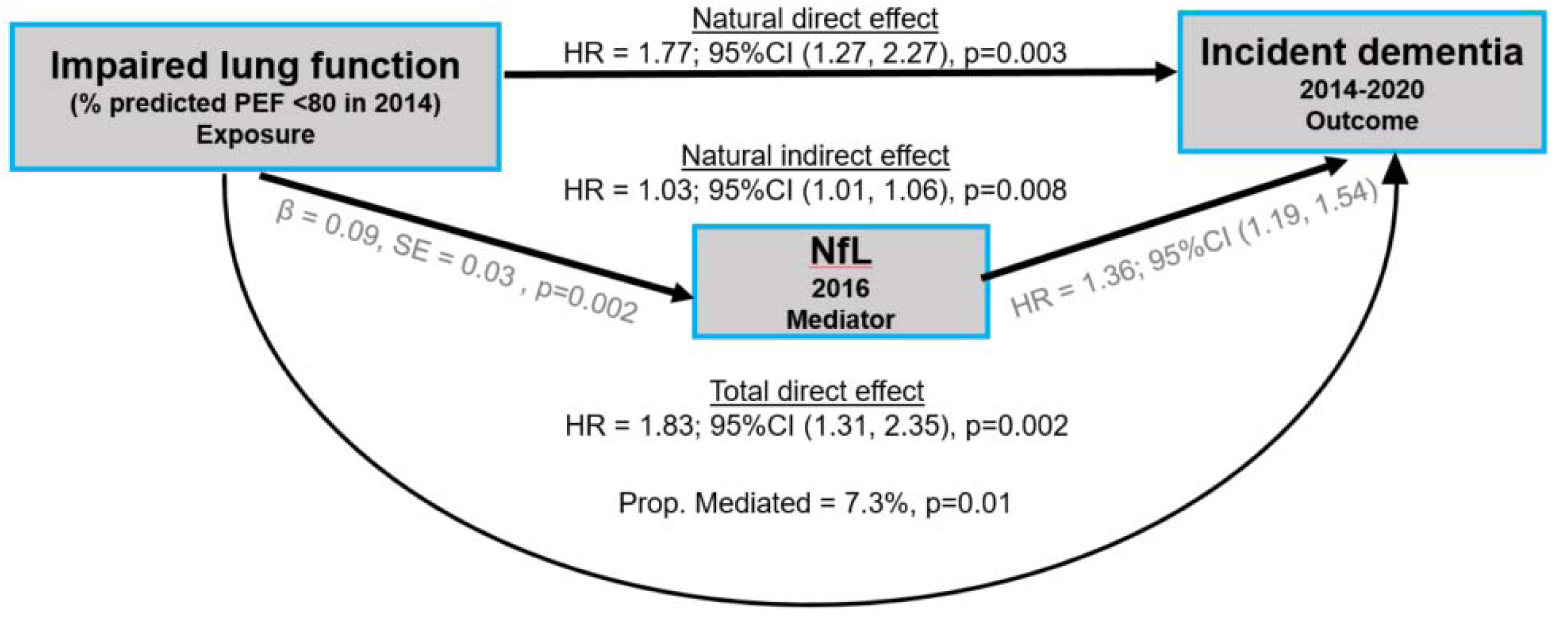
Causal mediation analysis of the association between decline in % predicted PEF and incident dementia mediated by plasma NfL. The path diagram of the causal mediation model with a three-variable system. In Path A, Impaired lung function ILF (exposure) has a significantly positive relationship with NfL (mediator) in a multivariable linear regression model. In Path B, ILF (exposure) was shown as an independent predictor for incident dementia (outcome) in the multivariable Cox hazards regression model without NfL. In Path C, both ILF (exposure) and NfL (mediator) remained significant to predict incident dementia using the multivariable causal mediation model with NfL partially mediate the association between ILF and incident dementia. HR: hazard ratio; CI: confidence interval, β: regression coefficient.

**Table 2A:** Association between percent predicted PEF in 2012/2014 (baseline) and blood biomarkers of AD in 2016.

| AD biomarkers | Q1 | Q2<br>$\beta$ (SE), p value | Q3<br>$\beta$ (SE), p value | Q4<br>$\beta$ (SE), p value | 1 SD unit<br>$\beta$ (SE), p value |
| --- | --- | --- | --- | --- | --- |
| Aβ 42/40 ratio ~ % pred PEF | 0.04 (0.05), 0.42 | 0.03 (0.05), 0.48 | 0.008 (0.04), 0.86 | Reference | -0.01 (0.02), 0.4600 |
| p-Tau 181 ~ % pred PEF | 0.15 (0.04), 0.0002 | 0.09 (0.04), 0.03 | 0.11 (0.04), 0.007 | Reference | -0.04 (0.01), 0.0040 |
| NfL ~ % pred PEF | 0.11 (0.03), 0.0006 | 0.09 (0.03), 0.005 | 0.02 (0.03), 0.63 | Reference | -0.05 (0.01), <.0001 |
| GFAP ~ % pred PEF | 0.10 (0.03), 0.004 | 0.09 (0.03), 0.008 | 0.07 (0.03), 0.03 | Reference | -0.04 (0.01), 0.001 |

When using impaired lung function (ILF; percent predicted PEF <80%) as a binary predictor, multivariable-adjusted linear regression models showed that individuals with ILF had significantly higher levels of p-Tau181 (β = 0.10, SE = 0.03, p = 0.004) and NfL (β = 0.09, SE = 0.03, p = 0.002), but no significant difference in GFAP (β = 0.04, SE = 0.03, p = 0.22) or Aβ42/40 ratio (β = 0.03, SE = 0.04, p = 0.52) compared to those with normal lung function (Table 2B). Beta coefficients represent standardized mean differences in biomarker levels between ILF and normal PEF groups.

**Table 2B:**
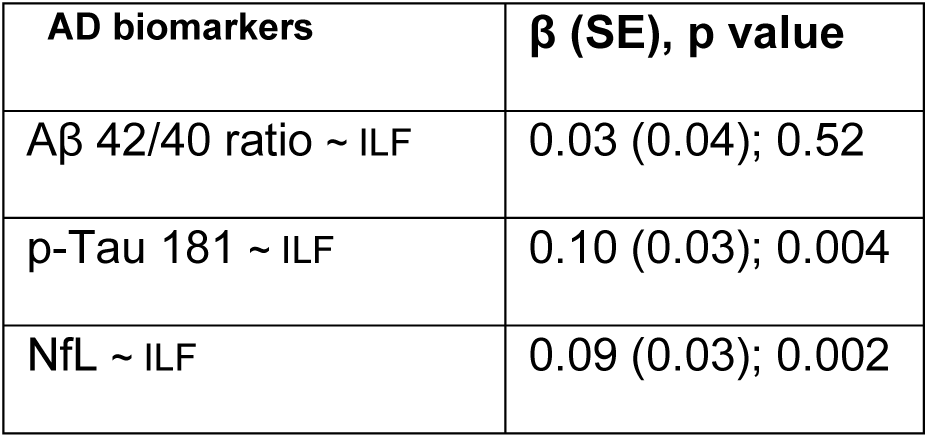
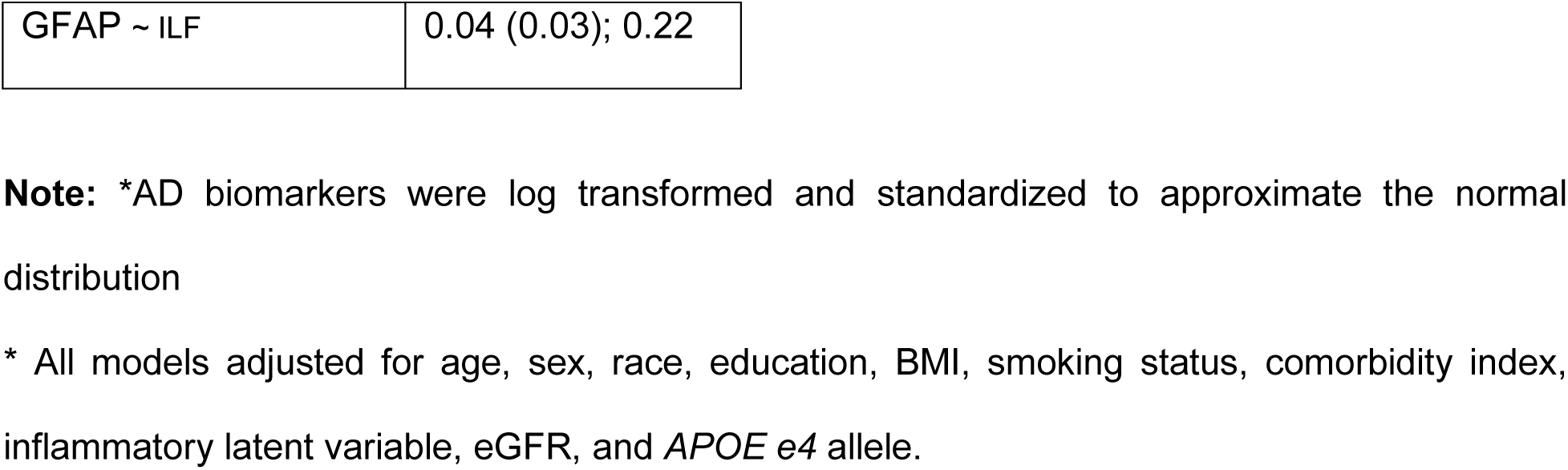
Association between impaired lung function in 2014 and blood biomarkers of AD in 2016.

Additional adjustment for systolic blood pressure and blood glucose levels measured in 2016 did not materially alter the associations, indicating that the observed links between impaired lung function and higher levels of p-Tau181 and NfL are robust.

### Decline in lung function and AD biomarkers

A greater decline in % predicted PEF from 2012/2014 to 2016/2018 was significantly associated with higher levels of plasma NfL (β (SE) = 0.07 (0.01), p<.0001) and GFAP (β, (SE) = 0.05 (0.01), p=0.0002) and serum p-Tau 181 (β(SE) = 0.05 (0.02), p= 0.0020) at 2016 after adjusting for baseline lung function and covariates (Table 2C). When modeling the decline in PEF as quartiles, individuals in the greatest decline group (Q4) had significantly higher levels of NfL (β = 0.18, SE = 0.03, p < 0.0001), GFAP (β = 0.09, SE = 0.03, p = 0.008), and p-Tau181 (β = 0.11, SE = 0.04, p = 0.009), compared to those with the least decline (Q1).

**Table 2C:** Association of decline in % predicted PEF from 2014 to 2016 as quartile and continuous variable with blood biomarkers of AD in 2016.

| AD biomarkers | Q1 | Q2<br>$\beta$ (SE), p value | Q3<br>$\beta$ (SE), p value | Q4<br>$\beta$ (SE), p value | 1 SD unit<br>$\beta$ (SE), p value |
| --- | --- | --- | --- | --- | --- |
| Aβ 42/40 ratio | Reference | -0.02 (0.05), 0.69 | -0.04 (0.05), 0.42 | -0.03 (0.05), 0.51 | -0.003 (0.02), 0.8500 |
| p-Tau 181 | Reference | -0.02 (0.04), 0.59 | 0.08 (0.04), 0.07 | 0.11 (0.04), 0.009 | 0.05 (0.02), 0.0020 |
| NfL | Reference | 0.05 (0.03), 0.12 | 0.08 (0.03), 0.02 | 0.18 (0.03), <.0001 | 0.07 (0.01), <.0001 |
| GFAP | Reference | -0.06 (0.03), 0.08 | 0.01 (0.03), 0.86 | 0.09 (0.03), 0.008 | 0.05 (0.01), 0.0002 |

Model – Adjusted for % predicted PEF in 2014 (baseline), age, sex, race, education, BMI, smoking status, comorbidity index 2014, inflammatory latent variable 2016, eGFR 2016, and *APOE e4* allele.

### Impaired lung function, AD biomarkers and risk of dementia

Individuals with impaired lung function (ILF: % predicted PEF < 80) had higher prevalence of dementia (OR=1.63, 95%CI = [1.04, 2.55]; p=0.0300) and CIND (OR=1.82, 95%CI = [1.47, 2.25]; p <.0001) in 2014 compared to those with normal PEF (% predicted PEF ≥ 80) after adjusting for age, sex, race, education, BMI, smoking status, comorbidity index in 2014 and inflammatory latent variable and eGFR in 2016, and *APOE e4* allele status. Among those participants without dementia in 2014 (n=3969, combined Normal and CIND), 6.9% (n=272) developed dementia over 6 years of follow-up. Individuals with ILF in 2014 had higher risk of developing dementia (HR=1.74, 95%CI = [1.34, 2.25]; p<.0001) compared to those with normal PEF after adjusting for all covariates. Including the AD biomarkers in the model as covariates reduced the strength of association between ILF and incident dementia (HR=1.67, 95%CI = [1.29, 2.17]; p=0.0001).

### AD biomarkers mediated the association between ILF and incident dementia over 6 years of follow up

Causal mediation analysis was performed to evaluate the role of AD biomarkers in the association between baseline impaired lung function (ILF) and incident dementia over a 6-year follow-up period among 3969 individuals without dementia in 2014 survey. Among the AD biomarkers associated with baseline ILF, plasma NfL and serum p-Tau 181 demonstrated partial mediation of this relationship in independent causal mediation models. Specifically, plasma NfL accounted for 7.3% of the association (p = 0.01) (Figure 2), while serum p-Tau 181 mediated 4.9% of the effect (p = 0.05, figure not shown) after accounting for baseline age, sex, race, BMI, smoking status, education, comorbidity index and *APOE e4*.

In a sensitivity analysis using dementia follow-up from 2016, we evaluated the mediation effect of NfL. The estimated proportion of the effect mediated increased slightly from 7.3% to 8.1%, although its statistical significance was attenuated (p-value increased from 0.01 to 0.07). Despite this, both the natural direct effect (OR = 1.55, p = 0.049) and the natural indirect effect (OR = 1.03, p = 0.044) remained statistically significant.

## DISCUSSION

In this study of older adults from the Health and Retirement Study, impaired lung function was associated with elevated levels of key blood biomarkers of AD including NfL and p-Tau 181 and increased risk of developing dementia over a 6-year follow-up period. To our knowledge, this is the first study to establish an association between impaired lung function and circulating AD protein biomarkers. Notably, we found that plasma NfL and serum p-Tau 181 partially mediated the association between baseline impaired lung function and future risk of dementia, suggesting a potential neurodegenerative pathway linking respiratory dysfunction to cognitive decline.

Blood biomarkers of AD/ADRD have gained significant attention in recent years due to their potential clinical utility in early identification and risk classification for neurodegeneration, dementia, and ultimately, Alzheimer’s disease^13,19^. Previous studies of AD biomarkers demonstrated that the cardiovascular and metabolic risk factors including BMI, renal function and vascular risk factors such as hypertension and diabetes affect the distribution of levels of AD biomarkers in blood^34,20,35^. Our study marks the first attempt to investigate the effect of lung function on blood biomarkers of AD. We demonstrated that lower baseline PEF is associated with higher levels of NfL and p-Tau 181 after two-four years of follow-up. Additionally, a greater decline in percent predicted PEF over a two-year period was significantly associated with elevated levels of NfL, p-Tau181, and GFAP, suggesting that deterioration in respiratory health may contribute to neurodegenerative processes. These associations were stronger and more consistent than those observed with lung function measured at a single time point, highlighting the importance of assessing change in lung function over time rather than relying solely on cross-sectional measures. These results suggest that accelerated decline in pulmonary function over a short period is linked to higher levels of biomarkers reflecting neurodegeneration and astrocytic activation, reinforcing the potential role of impaired respiratory health in early AD-related pathological processes. Prior research in cohort studies has established a link between impaired lung function and neuropathological changes, such as reduced brain volume and increased white matter lesions, suggesting potential mechanisms through which respiratory health may influence future cognitive decline.^36–38^ Our findings extend this evidence by demonstrating a link between impaired lung function and elevated levels of blood biomarkers of neuropathology, suggesting that neurodegenerative pathways linking impaired respiratory health and higher risk of dementia. Consistent with our findings of impaired lung function and elevated levels of AD biomarkers, a meta-analyses showed that reduced levels of FEV1 and FVC were significantly associated with diminished values of neuroimaging markers of AD (brain volume, gray matter volume, hippocampal volume, and higher volume of white matter hyper intensities)^11^. A large longitudinal study in the UK Biobank also established association between restrictive and obstructive impairment in lung function and all-cause dementia and brain MRI structural features of dementia^2^.

We found that baseline impaired lung function is associated with higher odds of having CIND and dementia. In our study, individuals with impaired lung function (PEF < 80%) had a 74% higher risk of developing dementia over a six-year follow-up period. Our findings are consistent with several reports of an association between better lung function and reduced dementia rate in other cohort studies^3,6,10^. Investigations in a younger cohort in the ARIC study with a longer follow-up also showed that individuals with impaired baseline lung function and restrictive/obstructive lung diseases have higher odds of cognitive impairment and dementia in later life^8,9^. Lutsey et al.^9^ reported that restrictive lung diseases, including idiopathic pulmonary fibrosis, were associated with a 58% increased risk of dementia or mild cognitive impairment (MCI), while obstructive lung diseases, such as COPD, were linked to a 33% higher risk. Another recent study in the ARIC cohort by Shrestha et al. with extended follow-up data reported that better lung function— measured by FEV1, FVC, and FEV1/FVC ratio—was associated with a slower cognitive decline across multiple domains and reduced dementia rate^4^. Additionally, prospective analyses from the CARDIA study, which followed participants from young adulthood to midlife, demonstrated that cumulative pulmonary function (FEV₁ and FVC measured repeatedly over 20 years) was associated with midlife cognitive performance. Specifically, cumulative FEV₁ and FVC were linked to better executive function (Stroop test) and psychomotor speed/attention (Digit Symbol Substitution Test (DSST)), even after adjusting for age, sex, race, smoking, and comorbidities. Notably, cumulative FEV₁ also showed a marginal association with verbal memory (RAVLT), suggesting lung health may differentially impact cognitive domains^1^. The Rotterdam Study highlights preserved ratio impaired spirometry (PRISm (FEV1/FVC≥70% and FEV1 < 80% predicted)) as underrecognized risk factors for dementia, independent of COPD^6^. They found that participants with FVC % predicted values in the lowest quartile compared to those in the highest quartile were at increased risk of all cause dementia (adjusted HR = 2.28; 95% CI = 1.31-3.98) and AD (HR = 2.13; 95% CI= 1.13–4.02), but no significant association was observed between FEV1 and FEV1/FVC ratio with incident all cause dementia or AD. These findings highlight that early-life impaired lung health, particularly restrictive lung function, increases susceptibility to cognitive impairment and dementia.

We demonstrated, for the first time, that plasma neurofilament light (NfL) and serum phosphorylated tau 181 (p-Tau 181) were identified as partial mediators, accounting for 7.3% and 5% of the association between impaired lung function and dementia risk, respectively, suggesting a potential biological pathway linking ILF to neurodegeneration. Though AD biomarkers measured concurrently when follow-up of dementia started. We performed sensitivity analysis following up participants after AD biomarker measures and observed that impaired lung function associated with incident dementia and NfL moderately mediated the association. These findings add to the growing body of evidence linking respiratory health to cognitive decline. A study examining the correlation between physical activity, serum NfL concentration, and cognitive decline found that participants with high levels of serum NfL who engaged in medium and high physical activity had a slower rate of cognitive decline compared to those with low physical activity^39^. This might suggest the potential influence of physical activity on improved lung function in mitigating the impact of Alzheimer’s disease pathology on cognitive function. Also, previous studies indicate that chronic hypoxia from respiratory illnesses such as COPD and sleep apnea can cause cognitive deficits, affecting attention, memory, and executive function^40^. This evidence highlights the need for clinical assessment of patients with lung function decline or COPD who have symptoms of neurodegeneration^41^. Evidence from a recent study on COVID-19 patients showed that higher GFAP levels at follow-up were associated with mild cognitive dysfunction. Since COVID-19 primarily affects respiratory function, its long-term impact on neuroinflammation and neurodegeneration has raised concerns about its potential role in Alzheimer’s disease (AD) development^42,43^. In our study, we observed that AD-related biomarkers—particularly those reflecting general neurodegeneration—partially mediated the relationship between impaired lung function and increased dementia risk in older adults. These findings are consistent with a hypothesized pathway in which lung impairment contributes to neurodegenerative processes, possibly through mechanisms involving hypoxia and systemic inflammation^20,44^. However, the complexity of these interactions suggests that additional factors may be involved, warranting further investigation to fully understand the underlying mechanisms. These findings highlight the importance of early detection of cognitive impairment through blood-based biomarkers in individuals with impaired lung function.

### Strengths and limitations

A major strength of this study is the availability of repeated measures of lung function, and cognitive function in a nationally representative sample of older adults, along with interim AD biomarker assessments. Additionally, the study enhances the robustness of the findings by effectively controlling for multiple confounding variables associated with both AD protein biomarkers and lung function. However, there are several limitations. First, only PEF was available as a measure of lung function, which may not fully capture respiratory impairment. Future studies incorporating more sensitive measures, such as FEV1 and FVC, are warranted. Second, AD biomarkers were measured at a single time point, limiting the ability to assess the longitudinal relationship between lung function decline and changes in AD biomarker levels. Third, in this study we investigated only four key AD-related biomarkers (Aβ42/40, p-Tau 181, GFAP, and NfL), which, while informative, do not capture the full spectrum of vascular dysfunction, neuroinflammation, or other potential pathways linking respiratory health to dementia. Future research should incorporate a broader panel of biomarkers, including markers of endothelial dysfunction, systemic inflammation, and cerebrovascular health, to better characterize the biological mechanisms underlying this association.

## Conclusion

In the Health and Retirement Study, impaired lung function was associated with elevated levels of key neuropathology biomarkers in blood, with NfL and p-Tau 181 partially mediating its association with risk of dementia. Our study findings highlight the importance of monitoring AD protein biomarkers in individuals with impaired respiratory health, which may help identify those at higher risk for cognitive decline and support timely interventions to mitigate neurodegenerative processes. These results warrant the need for further research to explore additional molecular biomarkers that mediate the association between impaired respiratory health and future risk of cognitive impairment and dementia.

## Data availability statement

We used the HRS publicly available datasets and sensitive biomarker data for this study analysis. This data can be found here: https://hrsdata.isr.umich.edu/data-products/public-survey-data and https://hrsdata.isr.umich.edu/dataproducts/sensitive-health and can be accessed by completing required data use agreement.

## Ethics statement

The venous blood study involving human samples was approved by University of Minnesota Institutional Review Board.

## Author contributions

SV: Conceptualization, formal analysis and methodology, writing – original draft, review and editing

EC: Data collection, Writing – critical review and editing.

JKK: Data development, Writing – critical review and editing.

JF: Data collection, Writing – critical review and editing.

DJ: Analysis methodology, Writing – critical review and editing.

WG: Analysis methodology, Writing – critical review and editing.

BT: Data collection, conceptualization, Writing – critical review and editing.

## References

1. Joyce BT, Chen X, Yaffe K, et al. Pulmonary Function in Midlife as a Predictor of Later-Life Cognition: The Coronary Artery Risk Development in Adults (CARDIA) Study. The Journals of Gerontology: Series A. 2022;77(12):2517–2523. doi:10.1093/gerona/glac026

2. Zhou L, Yang H, Zhang Y, et al. Association of impaired lung function with dementia, and brain magnetic resonance imaging indices: a large population-based longitudinal study. Age Ageing. Nov 2 2022;51(11)doi:10.1093/ageing/afac269

3. Donahue PT, Xue QL, Carlson MC. Peak Expiratory Flow Predicts Incident Dementia in a Representative Sample of U.S. Older Adults: The National Health and Aging Trends Study (NHATS). *The journals of gerontology Series A*, Biological sciences and medical sciences. Aug 2 2023;78(8):1427–1435. doi:10.1093/gerona/glac235

4. Shrestha S, Zhu X, London SJ, et al. Association of Lung Function With Cognitive Decline and Incident Dementia in the Atherosclerosis Risk in Communities Study. American Journal of Epidemiology. 2023;192(10):1637–1646. doi:10.1093/aje/kwad140

5. Xie W, Zheng F, Cai Y, et al. Reduced Lung Function and Cognitive Decline in Aging: A Longitudinal Cohort Study. Annals of the American Thoracic Society. Feb 2021;18(2):373–376. doi:10.1513/AnnalsATS.202009-1152RL

6. Xiao T, Wijnant SRA, Licher S, et al. Lung Function Impairment and the Risk of Incident Dementia: The Rotterdam Study. Journal of Alzheimer’s disease : JAD. 2021;82(2):621–630. doi:10.3233/jad-210162

7. Gilsanz P, Mayeda ER, Flatt J, Glymour MM, Quesenberry CP, Jr., Whitmer RA. Early Midlife Pulmonary Function and Dementia Risk. Alzheimer disease and associated disorders. Oct-Dec 2018;32(4):270–275. doi:10.1097/wad.0000000000000253

8. Pathan SS, Gottesman RF, Mosley TH, Knopman DS, Sharrett AR, Alonso A. Association of lung function with cognitive decline and dementia: the Atherosclerosis Risk in Communities (ARIC) Study. European journal of neurology. Jun 2011;18(6):888–98. doi:10.1111/j.1468-1331.2010.03340.x

9. Lutsey PL, Chen N, Mirabelli MC, et al. Impaired Lung Function, Lung Disease, and Risk of Incident Dementia. American journal of respiratory and critical care medicine. Jun 1 2019;199(11):1385–1396. doi:10.1164/rccm.201807-1220OC

10. Wang J, Dove A, Song R, et al. Poor pulmonary function is associated with mild cognitive impairment, its progression to dementia, and brain pathologies: A community-based cohort study. Alzheimer’s & Dementia. 2022/12/01 2022;18(12):2551–2559. 10.1002/alz.12625

11. Frenzel S, Bis JC, Gudmundsson EF, et al. Associations of Pulmonary Function with MRI Brain Volumes: A Coordinated Multi-Study Analysis. Journal of Alzheimer’s disease : JAD. 2022;90(3):1073–1083. doi:10.3233/jad-220667

12. Ashton NJ, Brum WS, Di Molfetta G, et al. Diagnostic Accuracy of a Plasma Phosphorylated Tau 217 Immunoassay for Alzheimer Disease Pathology. JAMA neurology. 2024;81(3):255–263. doi:10.1001/jamaneurol.2023.5319

13. Leuzy A, Mattsson-Carlgren N, Palmqvist S, Janelidze S, Dage JL, Hansson O. Blood-based biomarkers for Alzheimer’s disease. EMBO molecular medicine. Jan 11 2022;14(1):e14408. doi:10.15252/emmm.202114408

14. Mandal PK, Maroon JC, Garg A, et al. Blood Biomarkers in Alzheimer’s Disease. ACS chemical neuroscience. Nov 15 2023;14(22):3975–3978. doi:10.1021/acschemneuro.3c00641

15. Graff-Radford NR, Crook JE, Lucas J, et al. Association of Low Plasma Aβ42/Aβ40 Ratios With Increased Imminent Risk for Mild Cognitive Impairment and Alzheimer Disease. Archives of Neurology. 2007;64(3):354–362. doi:10.1001/archneur.64.3.354

16. Chatterjee P, Pedrini S, Doecke JD, et al. Plasma Aβ42/40 ratio, p-tau181, GFAP, and NfL across the Alzheimer’s disease continuum: A cross-sectional and longitudinal study in the AIBL cohort. Alzheimer’s & dementia : the journal of the Alzheimer’s Association. Apr 2023;19(4):1117–1134. doi:10.1002/alz.12724

17. Xiao Z, Wu W, Ma X, et al. Plasma p-tau217, p-tau181, and NfL as early indicators of dementia risk in a community cohort: The Shanghai Aging Study. Alzheimer’s & dementia (Amsterdam, Netherlands). Oct-Dec 2023;15(4):e12514. doi:10.1002/dad2.12514

18. Stevenson-Hoare J, Heslegrave A, Leonenko G, et al. Plasma biomarkers and genetics in the diagnosis and prediction of Alzheimer’s disease. Brain : a journal of neurology. Feb 13 2023;146(2):690–699. doi:10.1093/brain/awac128

19. Teunissen CE, Verberk IMW, Thijssen EH, et al. Blood-based biomarkers for Alzheimer’s disease: towards clinical implementation. The Lancet Neurology. Jan 2022;21(1):66–77. doi:10.1016/s1474-4422(21)00361-6

20. Rebelos E, Rissanen E, Bucci M, et al. Circulating neurofilament is linked with morbid obesity, renal function, and brain density. Sci Rep. May 12 2022;12(1):7841. doi:10.1038/s41598-022-11557-2

21. Hermesdorf M, Leppert D, Maceski A, et al. Longitudinal analyses of serum neurofilament light and associations with obesity indices and bioelectrical impedance parameters. Scientific Reports. 2022/09/23 2022;12(1):15863. doi:10.1038/s41598-022-20398-y

22. Yalachkov Y, Schäfer JH, Jakob J, et al. Effect of Estimated Blood Volume and Body Mass Index on GFAP and NfL Levels in the Serum and CSF of Patients With Multiple Sclerosis. Neurology(R) neuroimmunology & neuroinflammation. Jan 2023;10(1)doi:10.1212/nxi.0000000000200045

23. Sonnega A, Faul JD, Ofstedal MB, Langa KM, Phillips JW, Weir DR. Cohort Profile: the Health and Retirement Study (HRS). International journal of epidemiology. Apr 2014;43(2):576–85. doi:10.1093/ije/dyu067

24. Crimmins EM, Kim JK, Langa KM, Weir DR. Assessment of Cognition Using Surveys and Neuropsychological Assessment: The Health and Retirement Study and the Aging, Demographics, and Memory Study. The Journals of Gerontology: Series B. 2011;66B(suppl_1):i162–i171. doi:10.1093/geronb/gbr048

25. Crimmins E, Guyer H, Langa K, Ofstedal MB, Wallace R, Weir D. Documentation of physical measures, anthropometrics and blood pressure in the Health and Retirement Study. HRS Documentation Report DR-011. 2008;14(1-2):47–59.

26. Heeringa SG, Connor JH. Technical description of the Health and Retirement Survey sample design. Ann Arbor: University of Michigan. 1995;

27. Crimmins E, et al. Venus blood collection and assay protocol in the 2016 Health and Retirement Study 2016 Vensu Blood Study (VBS). 2017. Available at: https://hrsdata.isr.umich.edu/sites/default/files/documentation/data-descriptions/HRS2016VBSDD.pdf. Accessed June 2024.

28. DeVrieze BW, Goldin J, Giwa AO. Peak Flow Rate Measurement. StatPearls. StatPearls Publishing Copyright © 2025, StatPearls Publishing LLC.; 2025.

29. Hankinson JL, Odencrantz JR, Fedan KB. Spirometric Reference Values from a Sample of the General U.S. Population. American journal of respiratory and critical care medicine. 1999/01/01 1999;159(1):179–187. doi:10.1164/ajrccm.159.1.9712108

30. Panikkar D, Vivek S, Crimmins E, Faul J, Langa KM, Thyagarajan B. Pre-Analytical Variables Influencing Stability of Blood-Based Biomarkers of Neuropathology. Journal of Alzheimer’s disease : JAD. 2023;95(2):735–748. doi:10.3233/jad-230384

31. Meier HCS, Mitchell C, Karadimas T, Faul JD. Systemic inflammation and biological aging in the Health and Retirement Study. GeroScience. Dec 2023;45(6):3257–3265. doi:10.1007/s11357-023-00880-9

32. Faul J, Smith J, Zhao W. Health and retirement study: candidate gene and SNP data description. Health Retire Study, Univ Mich, Ann Arbor, MI http://hrsonline isr umich edu/sitedocs/genetics/candidategene/CandidateGeneSNPDataDescription Google Scholar Article Location. 2014;

33. Faul J, Smith J, Zhao W. Health and Retirement Study: Candidate genes for cognition/behavior. Ann Arbor, MI: University of Michigan. 2014;

34. Hoost SS, Brickman AM, Manly JJ, et al. Effects of Vascular Risk Factors on the Association of Blood-Based Biomarkers with Alzheimer’s Disease. Medical research archives. Sep 2023;11(9)doi:10.18103/mra.v11i9.4468

35. Pichet Binette A, Janelidze S, Cullen N, et al. Confounding factors of Alzheimer’s disease plasma biomarkers and their impact on clinical performance. Alzheimer’s & dementia : the journal of the Alzheimer’s Association. Apr 2023;19(4):1403–1414. doi:10.1002/alz.12787

36. Guo X, Pantoni L, Simoni M, et al. Midlife respiratory function related to white matter lesions and lacunar infarcts in late life: the Prospective Population Study of Women in Gothenburg, Sweden. Stroke. Jul 2006;37(7):1658–62. doi:10.1161/01.STR.0000226403.00963.af

37. Murray AD, Staff RT, Shenkin SD, Deary IJ, Starr JM, Whalley LJ. Brain White Matter Hyperintensities: Relative Importance of Vascular Risk Factors in Nondemented Elderly People. Radiology. 2005/10/01 2005;237(1):251–257. doi:10.1148/radiol.2371041496

38. Liao D, Higgins M, Bryan NR, et al. Lower pulmonary function and cerebral subclinical abnormalities detected by MRI: the Atherosclerosis Risk in Communities study. Chest. Jul 1999;116(1):150–6. doi:10.1378/chest.116.1.150

39. Desai P, Dhana K, DeCarli C, et al. Examination of Neurofilament Light Chain Serum Concentrations, Physical Activity, and Cognitive Decline in Older Adults. JAMA network open. Mar 1 2022;5(3):e223596. doi:10.1001/jamanetworkopen.2022.3596

40. Zheng GQ, Wang Y, Wang XT. Chronic hypoxia-hypercapnia influences cognitive function: a possible new model of cognitive dysfunction in chronic obstructive pulmonary disease. Medical hypotheses. 2008;71(1):111–3. doi:10.1016/j.mehy.2008.01.025

41. Areza-Fegyveres R, Kairalla RA, Carvalho CRR, Nitrini R. Cognition and chronic hypoxia in pulmonary diseases. Dementia & neuropsychologia. Jan-Mar 2010;4(1):14–22. doi:10.1590/s1980-57642010dn40100003

42. Miners S, Kehoe PG, Love S. Cognitive impact of COVID-19: looking beyond the short term. Alzheimer’s Research & Therapy. 2020/12/30 2020;12(1):170. doi:10.1186/s13195-020-00744-w

43. Ortiz GG, Velázquez-Brizuela IE, Ortiz-Velázquez GE, et al. Alzheimer’s Disease and SARS-CoV-2: Pathophysiological Analysis and Social Context. Brain sciences. Oct 18 2022;12(10)doi:10.3390/brainsci12101405

44. Kim KY, Shin KY, Chang KA. GFAP as a Potential Biomarker for Alzheimer’s Disease: A Systematic Review and Meta-Analysis. Cells. May 4 2023;12(9)doi:10.3390/cells12091309

